# Medical text prediction and suggestion using generative pre-trained transformer models with dental medical notes

**DOI:** 10.1101/2022.04.29.22274513

**Authors:** Joseph Sirriani, Emre Sezgin, Daniel Claman, Simon L Linwood

## Abstract

**Background:** Generative pre-trained transformer (GPT) models are one of the latest large pre-trained natural language processing (NLP) models, which enables model training with limited datasets, and reduces dependency on large datasets which are scarce and costly to establish and maintain. There is a rising interest to explore the use of GPT models in healthcare.

**Objective:** We investigate the performance of GPT-2 and GPT-Neo models for medical text prediction using 374,787 free-text dental notes.

**Methods:** We fine-tune pre-trained GPT-2 and GPT-Neo models for next word prediction on a dataset of over 374,000 manually written sections of dental clinical notes. Each model was trained on 80% of the dataset, validated on 10%, and tested on the remaining 10%. We report model performance in terms of next word prediction accuracy and loss. Additionally, we analyze the performance of the models on different types of prediction tokens for categories. We annotate each token in 100 randomly sampled notes by category (e.g. Names, Abbreviations, Clinical Terms, Punctuation, etc.) and compare the performance of each model by token category.

**Results:** Models present acceptable accuracy scores (GPT-2: 76%, GPT-Neo: 53%), and the GPT-2 model also performs better in manual evaluations, especially for names, abbreviations, and punctuation. The results suggest that pre-trained models have the potential to assist medical charting in the future. We share the lessons learned, insights, and suggestions for future implementations.

**Conclusion:** The results suggest that pre-trained models have the potential to assist medical charting in the future. Our study presented one of the first implementations of the GPT model used with medical notes.

## INTRODUCTION

Generative pre-trained transformer (GPT) models are one of the latest large pre-trained models designed to understand and produce natural language using a natural language understanding task performance architecture ^1^. Unlike many Artificial Intelligence/ Machine Learning (AI/ML) models, GPT has the capability of ‘few-shot,’ ‘one-shot,’ and ‘zero-shot’ learning, enabling model training with a limited dataset^2,3^. There has been increasing interest to use GPT models in the clinical domain^4,5^. Yet, they are limited to generating new text, without any focus on predicting subsequent text, which is one of the overarching capabilities of GPT in natural language generation. Current applications are towards creating synthetic clinical notes to increase data size^6^, generating summaries of medical trials^7^, and generating simplified clinical notes for patients^8^.

GPT-based text predictions have been implemented successfully in the industry (e.g. Microsoft text suggestions)^9^. Similar implementations could potentially help healthcare providers reduce the time spent on medical charting and improve clinician burn-out associated with time spent on medical charting^10^. A recent study shows that physicians spend approximately 16 minutes on Electronic Health Records (EHRs) for each patient, mostly in relation to creating notes and writing orders^11^. GPT is in a unique position to leverage AI/ML utilization in medical practice to support clinical tasks and medical charting without demanding significant resources to be trained and tested^4^.

In this study, we investigate the performance of 2 prominent open-access GPT models (GPT-2^12^ and GPT-Neo^13^) for medical text prediction using dental medical notes. Our research question is “To what extent can GPT models be utilized as an assistive tool (i.e. text suggestion) in writing medical notes?” We purposefully select dental notes as the study dataset, since dental notes structurally provide lesser language complexity in terms of formation and terminology as compared to medical notes from other specialties.

## MATERIALS AND METHODS

### Dataset

Our dataset consists of dental clinical notes at Nationwide Children’s Hospital’s data repository, containing over 1.2 million notes. We focus on 8 types of notes where most of the free-text medical information is collected: operative notes, examination notes, restorative notes, emergency notes, trauma notes, anesthesia post-evaluation notes, anesthesia pre-evaluation notes, and orthodontic clinical notes. The medical notes dataset is extracted from the EHR and stored as a string of text in our server. The notes include fields from structured sections and templates for each patient. In total, we have 374,787 notes consisting of different note types (**Table 1**). We separate the written free-text from the template text using regex rules^18^. We develop a regex rule for each note type to extract the purpose statement written by the dental clinician.

**Table 1.** A breakdown of the notes in the dataset by type.

| Note Type | Number of Notes |
| --- | --- |
| Operative Notes | 4390 |
| Exam Notes | 193,892 |
| Restorative Notes | 66,992 |
| Emergency Notes | 47,695 |
| Trauma Notes | 11,178 |
| Exam/Restore Notes | 3404 |
| Anesthesia Post Evaluation Notes | 16,716 |
| Anesthesia Pre-Evaluation Notes | 16,897 |
| Ortho Clinical Notes | 13,623 |
| Total | 374,787 |

### Prediction Model

GPT-2 is a large text generation model developed and released by OpenAI in 2019^19^. The large version of GPT-2 is utilized and contains 753 million parameters. GPT-2 is trained on the WebText dataset, a ∼40 GB collection of webpages from outbound links on Reddit^19^.

GPT-Neo is an open-sourced large text generation model developed by EleutherAI, and it is designed as a GPT-3-like model^20^, with billions of parameters. GPT-Neo is trained on the Pile, an 800GB-dataset of diverse texts from multiple sources^21^. We utilize the GPT-Neo version with 1.3 billion weights.

### Model Training

Utilized GPT-2 tokenizers contain 50,257 unique tokens. Each input is has a unique start-of-text token (“<|startoftext|>“) and an end-of-text token (“<|endoftext|>“) appended to their text. The GPT-2 model is trained on an NVidia V100 GPU, and the GPT-Neo model is trained on an NVidia A100 GPU. We limit the number of tokens considered by the model to 200 input tokens. This 200-token limit is selected by examining the distribution of the number of tokens of each note in the dataset, shown in **Figure 1**. Less than 0.4% of notes have 15 tokens or fewer, while over 70.1% of notes had between 16 and 50 tokens. In total, 94.6% of notes have fewer than 200 tokens total.

**Figure 1.**
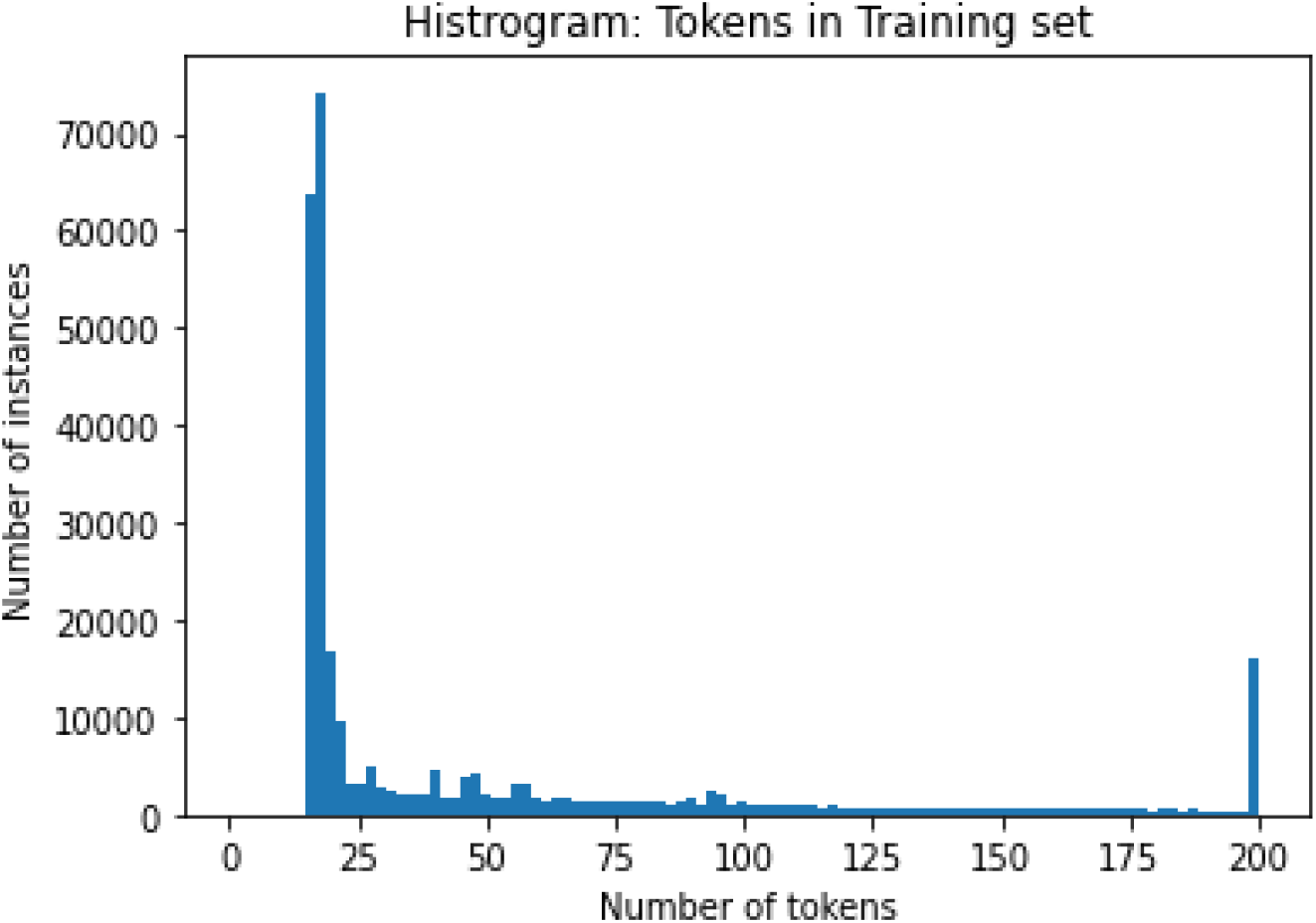
The distribution of the length of each note (in terms of tokens) in the training dataset, all instances with >200 tokens are binned into a single category.

GPT-2 and GPT-Neo are fine-tuned at default settings for the next token prediction for the manually written text of the dental clinical notes^12,13^. Both models are trained offline and have permissive MIT licenses. We use the Huggingface library^14^ to load both models and utilize a GPT-2 tokenizer. The dataset is divided into training-testing-validation datasets with an 80-10-10 split. Each model is fine-tuned for a maximum of 50 training epochs. Early-stopping is enabled with a tolerance of up to four epochs if the loss of the validation set did not improve. Both models are trained until the early stopping mechanism is triggered. The GPT models are trained for four epochs (GPT-2) and for 14 epochs (GPT-Neo).

### Ethics

This study is approved by the Institutional Review Board (IRB) of Nationwide Children’s Hospital (IRB No: 00000877).

## RESULTS

Model performance in terms of prediction loss^15^ (cross-entropy loss) and accuracy across the testing set are reported in **Table 2**. The testing accuracy represents the proportion of tokens that the model predicts correctly in the input sequence given the prior tokens. It is done without weighing in the padding token after the end-of-text token is encountered. In each model, the training loss scores are similar to validation loss and testing loss, indicating the model is not overfitting.^16^

**Table 2.** Quantitative performance metrics across models

| Model | Training Loss<br>(Average<br>Probability of<br>Correct Token) | Validation Loss<br>(Average<br>Probability of<br>Correct Token) | Testing Loss<br>(Average<br>Probability of<br>Correct Token) | Testing Accuracy<br>(all tokens) |
| --- | --- | --- | --- | --- |
| GPT-2 Large | **0.38 (0.684)** | **0.43 (0.651)** | **0.44 (0.644)** | **76%** |
| GPT-Neo 1.3B | 1.07 (0.343) | 1.07 (0.343) | 1.07 (0.343) | 53% |

### Token evaluation at sentence level

To understand the model performance in identifying accurately predicted words, the quality of the predictions, and differences between the models, we qualitatively review the outputs to characterize each model’s performance. We randomly sample 100 sentences from the testing set data and characterize each model’s performance on the different types of text. We annotate each token in the sentences into one of 10 categories: Capitalized words, numbers, punctuation/spaces, abbreviations, names, gender, encounter-related terms, diagnosis-related terms, other clinical terms, and regular words. **Table 3** presents the accuracy breakdown of each category. More specifically, the categories are as follows: 1) Capitalized Words (e.g. “PURPOSE”), 2) Normal English words, 3) Abbreviations (e.g. “Pt”, “yrs”), 4) Names, 5) Gender terms (e.g. “Male,” “Female”), 6) Diagnosis terms and patient condition (e.g. “gum pain”), 7) Punctuation and spaces, 8) Clinical Terms, 9) Numbers, and 10) Encounter related terms (e.g. “Postoperative,” “Sedation Evaluation”). For each model, we compare the token prediction accuracy for each token type. **Figure 2** visualizes an example prediction from GPT-2.

**Table 3.** Accuracy breakdown by each category

| **Description** | **Number of Tokens** | **GPT-2 Accuracy by category** | **GPT-Neo Accuracy by category** |
| --- | --- | --- | --- |
| Capitalized Words | 94 | 87.23% (82/94) | 86.17% (81/94) |
| Normal Words | 886 | 58.58% (519/886) | 37.02% (328/886) |
| Abbreviations | 149 | 61.07% (91/149) | 38.93% (58/149) |
| Names | 116 | 36.21% (42/116) | 0.86% (1/116) |
| Gender | 26 | 100.00% (26/26) | 46.15% (12/26) |
| Diagnosis/Patient Condition | 65 | 38.46% (25/65) | 0.00% (0/65) |
| Punctuation | 425 | 76.24% (324/425) | 42.12% (179/425) |
| Clinical Terms | 149 | 57.05% (85/149) | 10.07% (15/149) |
| Number | 57 | 40.35% (23/57) | 5.26% (3/57) |
| Encounter Related Term | 141 | 64.54% (91/141) | 47.52% (67/141) |
| **All Categories** | 2108 | 62.05% (1308/2108) | 35.29% (744/2108) |

**Figure 2.**
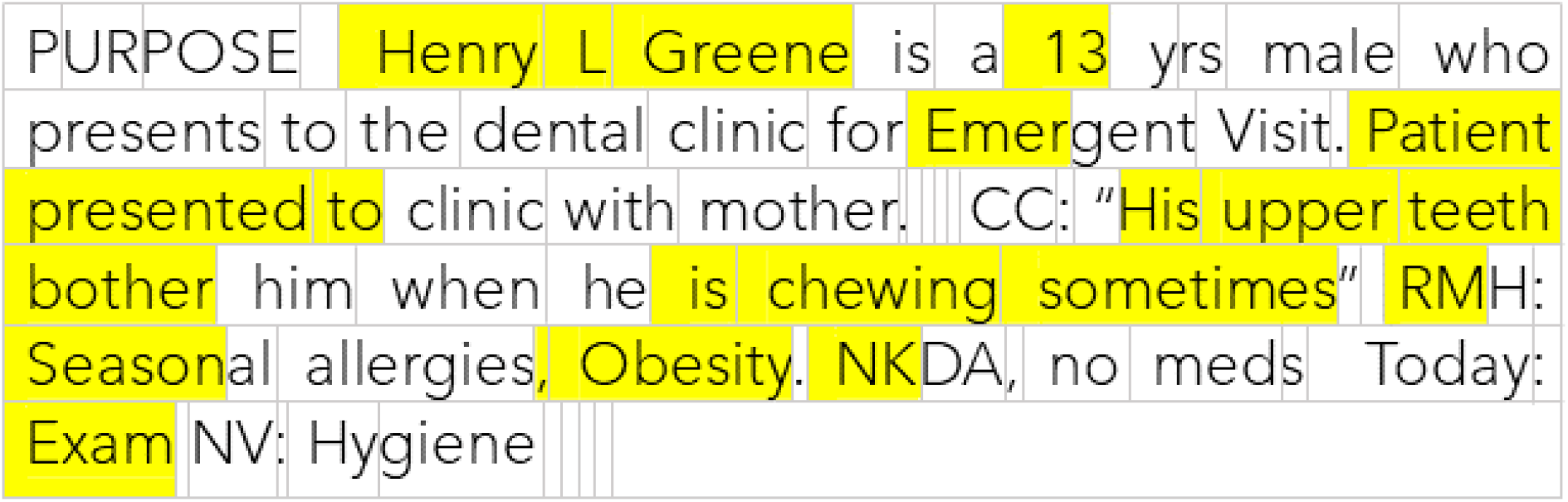
An example of a GPT-2 prediction on a simulated dental note. Each cell represents a token. A cell that is colored represents a missed prediction by the model. CC: chief complaint; RMH: recent medical history; NKDA: no known drug allergy; NV: next visit.

Overall, the GPT-2 model performs better than the GPT-Neo model by a 23% difference in terms of accuracy (GPT-2 score: 76%; GPT-Neo score: 53%). Based on the losses, we conclude that neither GPT-2 nor GPT-Neo experience significant overfitting, however, GPT-Neo has a substantially higher loss value than GPT-2. Considering GPT-Neo’s training with more epochs, it is likely that GPT-Neo is not able to effectively learn from the training data, possibly due to the small dataset, model size, and/or GPT-Neo being unable to retrain enough task-specific data due to its large size.

## DISCUSSION

Our results demonstrate the performance of text generation models for next word prediction on manually written dental notes. Overall GPT-2 performed competently predicting the next word in a note. However, it presents a number of blind spots when patient-specific information is missing. In terms of performance, the 76% accuracy for GPT-2 in our tasks is similar to performances for predicting 1-5 tokens for medical text simplification that is presented in an earlier study^17^. In the future, pre-trained model implementations can change the norms for clinical chartings, such as, replacing dot phrases or templates with intelligent guidance built-in to augment the free note-taking process (e.g. predicting note type, suggesting additional or missing entries).

For both models, a major source of error is predicting the patient-specific information in the note including patient name, age, gender, the reason for visit, and diagnosis. This is to be expected since the model has no prior information to base its prediction of the patient’s information. Structured patient-specific information fields from EHRs can be used to improve model-driven word prediction in the future. GPT-Neo has a lower accuracy in terms of clinical terms, abbreviations, and punctuations, which implies that it had more difficulty learning about the domain-specific terminology and formatting than GPT-2. Another large source of error is due to the abrupt changes in the topic in the dental notes. For example, a sentence about the procedure (e.g. patient presents for extraction of tooth) may be preceded by a sentence about medication, or about the general condition.

### Limitations and future works

Limitations of the study are the data source that came from a specific geographic region and is subject to standardized templates. In addition, the study dataset is only dental medical notes, not including other medical notes and domains. The lessons learned from this preliminary study could inform future works with more complex language use and complex tasks. Significant amounts of patient encounter information are expressed using programmatically generated text (e.g. templates), and thus are outside the scope of our investigation. These pieces of information may be contained in manually written notes at other organizations which could impact model performance.

We use GPT models which were minimally adjusted to the experiment. Future works could be extended to predicting longer sequences of notes, such as sentence completion within dental notes, and expanded to other medical domains. Currently, the fine-tuned GPT-2 has acceptable accuracy and frequently makes reasonable predictions. As our study is a preliminary effort, it does not provide clarity on the usefulness of the model for clinicians in practice. Future works could look at how clinicians would utilize a suggestion system by evaluating how often they accept suggestions. In addition, the new capability of word prediction could also introduce clinical errors when providers incorrectly accept the suggested words, which needs further evaluation. A suggestion to improve text prediction at the user side with GPT is training providers and developing charting guidelines with a GPT codebook developed to improve GPT model training and predictions.

## CONCLUSION

In this study, we investigate the performance of GPT-2 and GPT-Neo models for medical text prediction using free-text dental notes. Models present acceptable accuracy scores (GPT-2: 76%, GPT-Neo: 53%), and the GPT-2 model also performs better in manual evaluations. The results suggest that pre-trained models have the potential to assist medical charting in the future. Our study presented one of the first implementations of the GPT model used with medical notes.

## Data Availability

The dataset used in this study include private and sensitive information (e.g., medical records,
personal health information) which cannot be shared publicly.

## AUTHOR CONTRIBUTIONS

SL conceived the idea. All authors contributed to the design of the study. JS designed the experiments and conducted the analysis. DC supported retrieving the dataset. SL and DC supervised all parts of the study. JS and ES drafted the manuscript. All authors contributed the manuscript and approved the final version of it.

## DATA AVAILABILITY

The datasets used in this study include private and sensitive information (e.g., medical records, personal health information) which cannot be shared publicly. Please contact the corresponding author for your inquiries.

